# COVID-19 hospitalisation rates rise exponentially with age, inversely proportional to thymic T-cell production

**DOI:** 10.1101/2020.08.25.20181487

**Authors:** Sam Palmer, Nik Cunniffe, Ruairí Donnelly

## Abstract

Here we report that COVID-19 hospitalisation rates follow an exponential relationship with age, doubling for every 16 years of age or equivalently increasing by 4.5% per year of life (R^2^=0.98). This mirrors the well studied exponential decline of both thymus volume and T-cell production, which halve every 16 years. COVID-19 can therefore be added to the list of other diseases with this property, including those caused by MRSA, MERS-CoV, West Nile virus, Streptococcus Pneumonia and certain cancers, such as chronic myeloid leukemia and brain cancers. In addition, incidence of severe disease and mortality due to COVID-19 are both higher in men, consistent with the degree to which thymic involution (and the decrease in T-cell production with age) is more severe in men compared to women. Since these properties are shared with some non-contagious diseases, we hypothesised that the age-dependence does not come from social-mixing patterns, i.e. that the probability of hospitalisation *given infection* rises exponentially, doubling every 16 years. A Bayesian analysis of daily hospitalisations, incorporating contact matrices, found that this relationship holds for every age group except for the under 20s. While older adults have less contacts than young adults, our analysis suggests that there is an approximate cancellation between the effects of less contacts for the elderly and higher infectiousness due to a higher probability of developing severe disease. Our model fitting suggests under 20s have 49-75% additional immune protection beyond that predicted by strong thymus function alone, consistent with increased juvenile cross-immunity from other viruses. We found no evidence for differences between age groups in susceptibility to infection or infectiousness to others (given disease state), i.e. the only important factor in the age-dependence of hospitalisation rates is the probability of hospitalisation given infection. These findings suggest the existence of a T-cell exhaustion threshold, proportional to thymic output, and that clonal expansion of peripheral T-cells does not affect disease risk. The strikingly simple inverse relationship between risk and thymic T-cell output adds to the evidence that thymic involution is an important factor in the decline of the immune system with age and may also be an important clue in understanding disease progression, not just for COVID-19 but other diseases as well.

## Introduction

Epidemiological patterns in the incidence of a disease can provide insight into the mechanisms of disease progression^1–4^. The degradation of the adaptive immune system with age is already acknowledged to be a major risk factor for both infectious and non-infectious diseases and may play a role in understanding the emerging COVID-19 epidemic. Thymus volume, and the concomitant production of T-cells, decrease exponentially with age with a half-life of 16 years, or equivalently by 4.5% per year^5,6^ (Fig. S3A). These changes in the adaptive immune system contribute to less robust immune responses in elderly individuals^7^. In this paper, we analyse age and sex trends in national COVID-19 hospitalisation data, in order to investigate the role of immune function in the ongoing coronavirus pandemic.

COVID-19 disease progression can be characterised by three consecutive phases of increasing severity^8,9^. First, there are mild symptoms such as a dry cough, sore throat and fever. After this point the majority of cases will undergo spontaneous regression^10^. Second, some patients can develop viral pneumonia, requiring hospitalization^8^. The third stage, typically occurring three weeks after the onset of symptoms, is characterised by fibrosis^8^ and leads to life threatening symptoms^10–12^. COVID-19 patients often exhibit lymphopenia, i.e. extremely low blood T-cell levels, even in the first few days after the onset of symptoms, which is a predictor of disease progression and mortality^13,14^. Clinical trials are currently underway to test T-cell based immunotherapies^15,16^ and vaccines that elicit T-cell, as well as antibody, responses^17^. There is evidence that T-cells may be more effective than antibodies as exposed, asymptomatic individuals develop a robust T-cell response without (or before) a measurable humoral response^18^.

The relationship between COVID-19 risk and age has been extensively explored^19–22^ and age-stratified, contact-based, transmission models have accurately explained various aspects of the pandemic^20,21,23,24^. In particular, these studies have found that the risk of severe disease rises with age and is especially low for those under 20. Some studies suggest that non-adults are as likely to be infected as adults, but then have lower risk of disease progression^24^ while others find lower risk of both infection and disease progression in the under 20s^21,23^. While these studies have looked at COVID-19 risk and age, here we go further by relating these trends to thymic involution and T-cell production. This may lead to a mechanistic understanding of disease progression.

Several diseases have risk profiles that increase exponentially with age, doubling every 16 years, i.e. risk is proportional to e^0.044*t*^, where *t* is age, or equivalently increasing by about 4.5% per year^4^. These diseases are caused by a range of pathogens, from bacterial (MRSA, *S. Pneumonia*) to viral (West Nile virus, MERS-CoV^25^) and even include some cancers (chronic myeloid leukemia, heart and brain cancers). Since thymus volume and T-cell production both decrease with age exponentially, halving every 16 years^5^, disease risk is therefore inversely proportional to T-cell production for these diseases. A mechanistic model has been proposed to explain this inverse relationship, incorporating an immune escape threshold and stochastic fluctuations in antigen levels^4^. Furthermore, the sex bias in thymic involution (and T-cell production) also roughly matches the sex bias in disease risk, with men having approx. 1.3-1.5 times higher overall cancer and infectious disease risk^26–28^ and approx. 1.5±0.3 times lower T-cell production, as measured by T-cell receptor excision circles (TRECs), a proxy for thymic output^4,6^. As such, fundamental patterns in disease incidence with respect to both age and sex can be directly linked to differences in the adaptive immune system. We therefore tested to see if COVID-19 follows the same trend.

## Results

### COVID-19 hospitalisation rates

While data on confirmed cases can be highly variable and largely influenced by testing strategies, the data on hospitalisations, which is the focus of this paper, are relatively more reliable. Incidence of COVID-19 hospitalisations, in a number of countries, consistently doubles with every 16 years of age (R^2^=0.98 for top three countries, Fig. 1A). Meanwhile, the incidence of all confirmed cases (including mild or asymptomatic) appears roughly constant across adult ages (Fig. S1). One explanation that is consistent with the data is that exposure is approximately uniform for adult age groups and that after exposure, the probability of becoming hospitalised is proportional to e^0.044*t*^, where *t* is age. We will address the age-dependence of exposure in more detail by accounting for assortative social mixing as well as a range of additional age-dependent factors in our Bayesian model (see below).

**Fig. 1.**
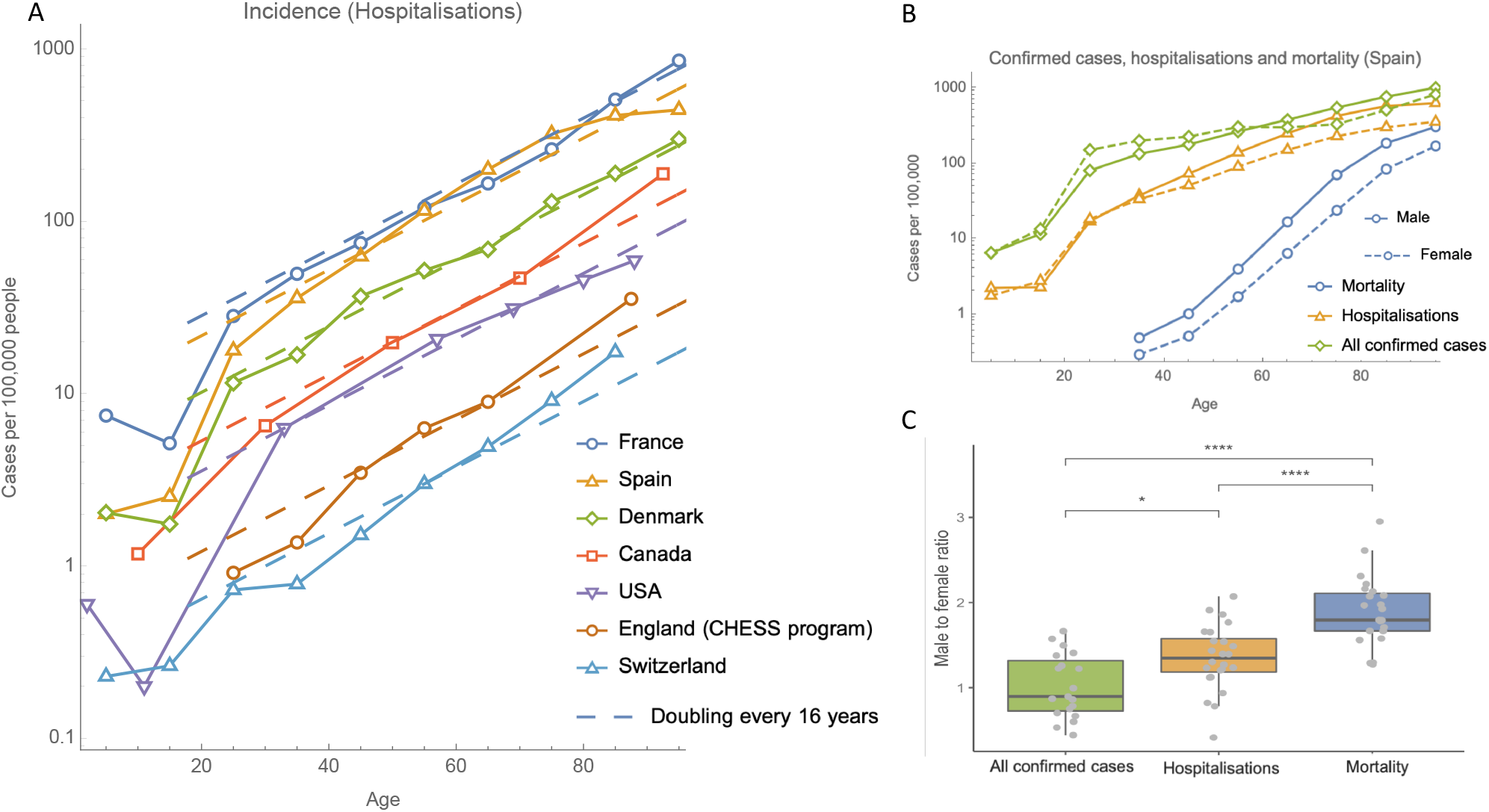
(A) For adults, incidence of COVID-19 hospitalisations rises exponentially with age, doubling with every 16 years of age. See table S3 for a full list of data sources. (B) Data from Spain on all confirmed cases, hospitalisations and mortality, from a single study early in the epidemic, shows a sex bias which increases with disease severity. (C) Boxplot showing male to female ratios for incidence, hospitalisation rates and mortality, across all age groups with non-zero entries, from the following countries: France, England, Wales and Spain.

There is a sex bias in COVID-19 risk, which increases with disease severity (Fig. 1C). This is similar to other diseases, including cancer, where men have 1.33 times the risk of hospitalisation and 1.89 times the risk of death^26,29^. The sex bias in COVID-19 is remarkably similar with a factor 1.35±0.4 for hospitalisation incidence and 1.9±0.4 for mortality (mean ± s.d. Fig. 1C). The slope of the logarithm of the COVID-19 mortality curve is over twice that of the hospitalisation curve, corresponding to an exponential with rate 0.109±0.005 years^−1^ (Fig. 1B). Another way of thinking about the sex bias would be to say that for both hospital incidence and mortality, men are effectively ∼6 years older than women in terms of risk. Other risk factors such as BMI can also be viewed similarly to give an individualised effective “Covid age”^30^. The increase in mortality with age may also be explained by comorbidities which increase with age, such as cardiovascular disease, which rises exponentially^31^ with a rate of 0.071±0.003 years^−1^. Since 0.071+0.044=0.115≈0.109, a simple model where the risk of COVID-19 mortality is proportional to risk of cardiovascular disease and inversely proportional to T-cell production would have the correct age-dependence. This would suggest that cardiovascular disease is a risk factor just for the stages in between hospitalisation and death.

### Bayesian model

Similar to other diseases, COVID-19 hospitalisation risk is relatively high for very young children (e.g. 0.6 cases per 100,000 for ages 0-4 *vs*. 0.2 cases per 100,000 for ages 5-17 in USA, Fig. 1A). Additionally, older children have a risk lower than expected based on the exponential increase with age we have identified (Fig. 1A). This is similar to MRSA and *S. Pneumonia* infection, but not West Nile virus (WNV) infection or cancers with similar exponential behaviour ^4^. Potential factors underlying the apparent low risk in juveniles include age-dependence in: 1) exposure (e.g. due to heterogeneous social mixing among age groups), 2) disease progression, 3) infection given exposure, and/or, 4) infectiousness to others. Throughout this paper we use the term ‘severe infection’ synonymously with hospitalisation and we categorise all infections as either mild or severe. In a preliminary analysis, we first incorporated contact matrices into a simple analytically-tractable Susceptible-Infected-Removed (SIR) model to predict the steady state of the age distribution of hospitalisations in France, with the assumption that the probability of severe disease given infection is proportional to e^0.044t^ (see supplementary materials, Fig. S3). This model suggested that age differences in social mixing could, in part, account for the relatively low hospitalisation of non-adults (Fig. S3). However, the other possible factors in low juvenile COVID-19 hospitalisation were not considered in this preliminary analysis.

To incorporate all relevant factors, and to rigorously test the hypothesis that the probability of hospitalisation given infection rises with age at the same rate as thymic involution, we conducted a more detailed analysis of age-dependence based on daily hospitalisation, recovery and death data. We focused on the single country France, for which an unusually comprehensive age distributed dataset is available^20^. All cases in the dataset are either biologically confirmed or present with a computed tomographic image highly suggestive of SARS-CoV-2 infection, and the dataset includes corrections for reporting delays^20^. We formulated an age-structured Bayesian SIR model of infection, partitioning the force of infection into that arising from contacts with mild and severely infected individuals, weighted by age-dependent contact matrices, as well as contact-independent (environmental) transmission. The model fitting exercise focused on inferring a posterior parameter distribution for the probability of severe disease given infection for each age cohort. In addition, posterior distributions were inferred for a range of secondary parameters (Table S2, parameters of the Bayesian analysis), including age-dependent transmissibility and susceptibility.

Our results reiterate that the probability of severe disease given infection increases exponentially with age, at a rate that is remarkably well matched by the rate of thymus decline for all age groups above 20 years (Fig. S4, all adult age groups have 95% credible intervals including the rate of thymus decline). In order to investigate the nature of juvenile deviation from this exponential relationship, we reformulated the analysis to allow deviations from an exponential increase (for the probability of severe disease given infection) for each age cohort (Fig. 2). The posterior parameter distribution for the exponential rate was found to match the rate of thymic degradation (95% CI:0.043-0.053 years^−1^, Fig. 2B). Only the juvenile age-cohort was found to significantly deviate from the exponential response (Fig. 2C), showing a level of additional protection to severe COVID-19 of between 49-75% (Table S1). Our sensitivity analysis allowed – within each age cohort – for deviation from uniform probability of infection given exposure, and, deviation from uniform infectiousness of infected individuals. For both of these we found that none of the age cohorts deviated significantly (in all cases 95% credible intervals included zero deviation, Fig. S5), allowing us to discount these potentially confounding factors.

**Fig. 2.**
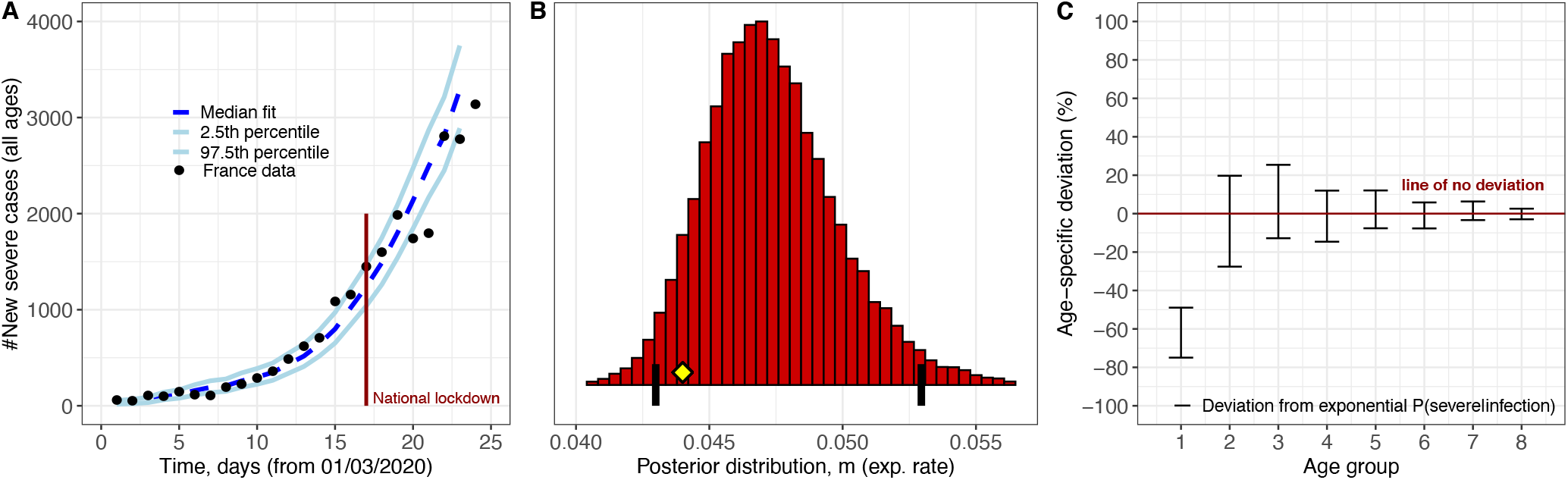
(A) Forward simulation of the French epidemic using the fitted parameters (B-C) produces a credible interval containing the French hospitalisation data up to day 24 (B) The 95% credible interval for the rate of age dependent exponential growth in hospitalisation probability includes the rate of thymus degradation (0.044 years^−1^, yellow diamond). Black vertical lines show the 2.5^th^ and 97.5^th^ percentiles. (C) The juvenile cohort has additional significant protection beyond what is predicted by their stronger thymus function (red interval is separated from the zero deviation line for juveniles only). See supplementary information section ‘Bayesian modelling’ for full description of methods.

The low susceptibility to severe disease given infection in non-adults may be due to cross-protection from other coronaviruses^8,32–34^, or even non-specific protection from other respiratory viruses^35^, which occur more frequently in non-adults compared to adults^36^. Our estimate of 49-75% protection ties in with a study which found SARS-Cov-2 reactive antibodies in approximately 60% of unexposed individuals aged 6-16 and only 6% in adults^37^. There is also evidence of unexposed individuals having SARS-CoV-2 reactive CD4+ T-cells^38^. Another possible explanation for the low risk in non-adults might come from some intrinsic feature of the immune system. For example, we speculate that the cause may be related to the high risk of T-lymphoblastic leukemia for ages 5-20 (see ref.^4^ Supp. Fig. 5).

## Discussion

Although we have demonstrated a clear relationship between the probability of severe disease and age, it is possible that the relationship is due, in part, to alternative physical processes other than T-cell production, such as age-related changes in the bone marrow, spleen or lymph nodes. Bone marrow also shrinks with age, but at a rate that is substantially slower than the thymus^39^. Further experiments are needed to determine the degree of causation between T-cell production and disease risk, for example by measuring TRECs and performing a prospective study or quantifying the increase in risk in thymectomized individuals^40^. Furthermore, a mechanism for why the probability of hospitalisation is inversely proportional to T-cell production is currently lacking. One possible model features stochastic fluctuations in the number of infected cells and an immune escape threshold which is proportional to T-cell production^4^. This model has the added benefit that it can also explain most of the other (non-exponential) relationships between risk and age seen in various cancer types^4^.

Chronic myeloid leukemia (CML) is a type of cancer with an age-dependence remarkably similar to COVID-19. In both diseases the risk of hospitalisation rises exponentially, inversely proportional to T-cell production^4^, with sex bias ratios of 1.35±0.4 for COVID-19 and 1.35±0.3 for CML. The mortality risk profiles are also similar (exponential rates: 0.109±0.005 years^−1^ for COVID-19 and 0.103±0.007 years^−1^ for CML, sex bias ratios:1.9±0.4 for COVID-19 and 1.8±0.6 for CML, Fig. S2). CML is characterised by a single genomic feature, a chromosomal translocation known as the Philadelphia chromosome. This suggests that the probabilities of Philadelphia chromosome formation and COVID-19 infection are approximately age-independent, but that the probabilities of subsequent hospitalisation are T-cell dependent. A good candidate for a potential mechanism involves the phenomenon that increased antigenic load can lead to T-cell exhaustion, characterised by low effector function and clone-specific depletion^41^. T-cell exhaustion is a factor in both cancer and infectious diseases, including COVID-19^42,43^, where it has even been shown to be a predictor of mortality^44^. As T-cell production decreases with age, this may lead to an increase in the probability for T-cell exhaustion. In support of this hypothesis, low precursor T-cell numbers have been shown to lead to T-cell exhaustion and disease progression in a mouse cancer model^45^. More specifically, we predict a step in disease progression with a probability exactly inversely proportional to the number of precursor T-cells.

When looking at sex biases for COVID-19 hospitalisation and mortality (Fig. 1C) we found factors of 1.35 and 1.9 respectively. We can speculate that since 1.35^2^≈1.9, this might be an indication that among the steps of disease progression, there could be two T-cell dependent steps, one pre-hospitalisation and one post-hospitalisation. This would imply that the risk of death would involve two factors of e^0.044t^ and therefore mortality would increase at least as fast as e^0.088t^. The log-slope of the mortality curve being 0.109±0.005 years^−1^ is consistent with this hypothesis. One feature of post-hospitalisation disease progression is an IL-6 driven cytokine storm^46^, which has been related to T-cell dysfunction in a mouse model^47^. These T-cell dysfunction-related cytokine storms were attenuated by nicotinamide adenine dinucleotide precursors and blocking of TNFα signalling.

## Conclusion

Here we have shown that risk of COVID-19 hospitalisation rises exponentially with age, inversely proportional to T-cell production, in a similar way to several other diseases. Consistently, the sex bias in disease risk also fits this trend. These features suggest that the risk of hospitalisation is related to an immune deficiency, rather than an immunopathology. In contrast, long-COVID follows patterns similar to autoimmune diseases, with middle-aged women having the highest risk^48^. In addition, we found that the under-20 age group benefits from additional protection from severe disease by a factor similar to the prevalence of SARS-CoV-2 cross-reactive antibodies. Our mathematical model suggests that the age dependence of hospitalisation rates does not arise from differences in social-mixing patterns, but rather from the probability of hospitalisation given infection. The model could be easily extended to assess which age-groups and socioeconomic-groups would be most valuable to vaccinate and therefore to optimise vaccination strategies.

These findings add to the growing evidence that thymic involution is a major component of immunosenescence and that restoring thymus function may be an effective preventative measure for many common diseases. Additionally, our understanding of host-pathogen dynamics is not complete. There is currently no detailed mechanistic model to explain why the probability of hospitalisation would be proportional to the reciprocal of thymic T-cell production, for COVID-19 or any other disease. We hope that these findings will be an important clue in understanding the precise mechanisms involved in disease progression.

## Data Availability

Additional code can be found at https://github.com/samIndeed/COVID

## Acknowledgments

We would like to thank Mark Coles, Eamon Gaffney, Jon Chapman, Philip Maini and Marc-Andre Rousseau for comments. In particular we would like to thank Thea Newman for help with an early draft and for the idea to think of risk in terms of effective ages. We acknowledge financial support from Wellcome Trust (Grant ID 211944/Z/18/Z).

## Author Contributions

SP developed the initial idea, collected data, did statistical analysis and drafted the manuscript. NC contributed to interpretation of the data and results and to critically revising the manuscript. RD collected data, developed Bayesian modelling, contributed to interpretation of the data and results and to critically revising the manuscript. All authors gave final approval for publication and agree to be held accountable for the work performed therein.

## Data and materials availability

All data and code is available in the main text or the supplementary materials or at https://github.com/samIndeed/COVID.

## Supplementary Materials

### Methods

In the ‘Analytical models’ section below we include technical details on the deterministic model used to compare theory with data in Figure S3. In the ‘Bayesian models’ section below we include technical details on the statistical model used to generate the results displayed in Figure 2, S4-S6 and Table S1.

#### Analytical models

We consider three models: one with age-independent spreading, one with contact-based spreading and no transmission from those infected with mild symptoms and one with contact-based spreading where those with mild symptoms are as contagious as those with severe symptoms. Throughout this paper we use the term ‘severe’ to correspond to hospitalisations. The first model is a simple model where we assume that transmission and exposure are age-independent and that the probability of subsequent severe infection is proportional to e^0.044*a*^, where *a* is age. This model predicts hospitalisation rates to rise as a pure exponential with exponential rate 0.044 years^−1^.

In our contact based models, we assume that risk of coronavirus infection is of the form

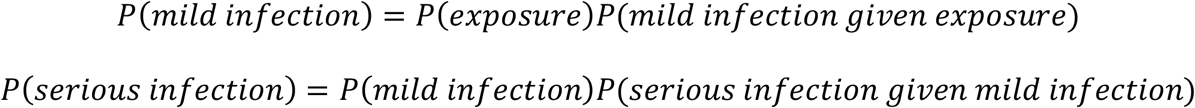

and that *P*(*mild infection given exposure*) is age-independent while *P*(*serious infection given mild infection*) is proportional to e^0.044*a*^. We also assume that *P*(*exposure*) for someone aged *i* is proportional to the number of people infected at age *j* times the amount of contact between age *i* and age *j* (as measured by a contact matrix which we will call *C*). Then the number of infected people can be modelled as a discrete time Markov process. In what follows, we consider only new cases each time step, which is equivalent to having one time step being the length of time someone is infectious for. We also make the approximation that the number of susceptibles is much larger than the number of infected and recovered/dead.

If individuals with only mild symptoms do not transmit, we can ignore them in our model. If the number of people severely infected at time *t*, for each age group, is the vector ***n***_*t*_ then we have (up to a constant of proportionality setting how fast the epidemic grows)

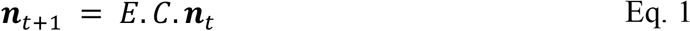

where *C* is the contact matrix and *E* is a diagonal matrix with e^0.044*a*^ on the diagonal. Since all elements of *C* are greater than zero, the Markov chain is strongly connected and therefore, regardless of initial conditions, ***n***_*t*_ will be dominated by the term

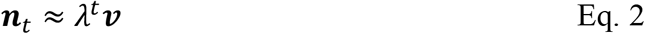

where ***v*** is the eigenvector of *E*.*C* with the largest eigenvalue, *λ*. Therefore the normalised age-distribution of new (and cumulative) severe-symptom cases will converge to ***v***. The predicted incidence will then be proportional to ***v*** divided by the number of people in each age group, which comes from the population age distribution.

If the mild-symptom individuals are as contagious as the severe-symptom individuals, we let the number of infected (both mild and severe) individuals at time *t*, for each age group, be the vector ***m***_*t*_. Then, up to a constant, we have

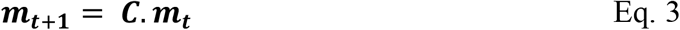

where *C* is the contact matrix. Again, since the Markov chain is strongly connected, regardless of initial conditions, ***m***_*t*_ will be dominated by the term

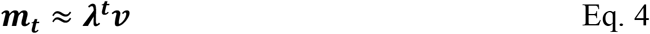

where ***v*** is now the eigenvector of *C* with the largest eigenvalue *λ*. The age-distribution of severe-symptom cases would then be proportional to *E*.***v***. The overall constant is then an arbitrary fitting parameter, which we fit to the actual incidence data.

#### Bayesian models

C we describe a model focusing on severely infected individuals. This model is the basis of the Bayesian analysis, i.e., where the unknown parameters of the statistical model are fit to data using MCMC methods. In what follows, the unknown, i.e., fitted, parameters are highlighted in bold.

The number of severe infections (hospitalisations) at time *t+1* in age group *i* is denoted by *Sev*^*t*+1^_*i*_. Similarly, *Mild*^*t*+1^_*i*_ denotes the number of mild infections (i.e., infections that are not severe). The number of severe infections that arise on a particular day in a given age group has the distribution:

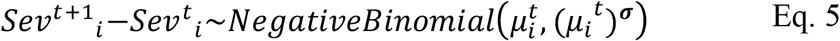

where

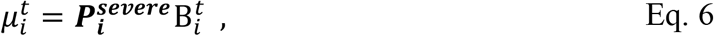

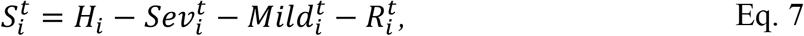

and

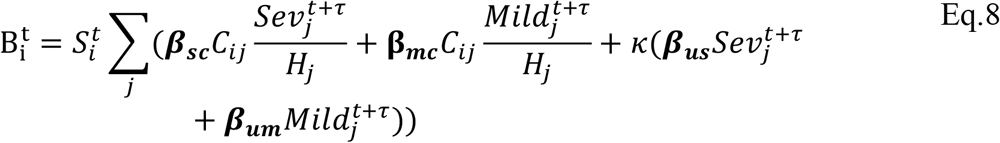

In Eq.s 5-8 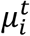 is the mean number of new severe infections produced in age group *i* on day *t*, 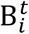 is the force of infection for individuals in age group *i* on day *t* that are susceptible to infection (denoted 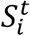). In addition, 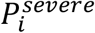 represents the probability of severe disease given infection in age group *i, β*_*sc*_ and *β*_*mc*_ represent the contact-dependent transmission rates from severe and mildly infected individuals respectively. *β*_*su*_ and *β*_*mu*_ represent the contact-independent, i.e., environmental transmission rates from severe and mildly infected individuals respectively. The parameter *κ* is a simple scaling parameter which does not alter the analysis that is included in order to ensure that the contact-dependent and contact-independent transmission rates are on a comparable scale (i.e., to allow fitting of the ratio of these terms; 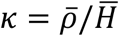 where *ρ*_*i*_ is the mean number of contacts that a particular age group, indexed *i*, makes with other age groups, 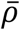 is the mean of ***ρ*** and H is the mean of ***H***). Finally, τ represents the average delay between the force of infection and the time when a patient is admitted to hospital, which we take to be 0 days with several other values (representing *τ* > 0) chosen in the sensitivity analysis (see *outline points* below for further details). See Table S2 for a complete set of fitted parameter descriptions. Thus we model the infection process as a negative binomial distribution. This accounts for the variation in count data associated with the occurrence of individual cases in a given age group on a given day, while allowing for potential over-dispersion in count data which may arise, for instance, through the aggregation of French regions, having distinct epidemics. In addition, we model the occurrence of severely infected individuals in an age group given a set number of daily infections in that age group, as a binomial distribution (taken together the binomial and negative binomial distributions result in an overall negative binomial distribution for the occurrence of severe cases, Eq. 5).

The following outline points clarify the choices made in formulating the model:

- In our model, the number of new hospitalisations at time *t* depends on the number of people in hospital at time *t*+τ. Our choice of τ=0 comes from assuming a typical patient would be admitted to hospital ∼10 days ^48^ after infection and stay in hospital for ∼10 days ^48^. Therefore hospital admissions at time *t* would depend on prevalence at *t*-10 and prevalence at *t*-10 would lead to hospital occupancy anywhere from *t*-10 to *t*+10. Taking the key points from this interval gives *τ* = 0, *τ* = 5 and *τ* = 10. We chose to match the simplicity of our approach to the simplicity of our purpose and hence assumed that τ=0 in our main analysis. For comprehensiveness, however, we assumed the other key points from the interval in our sensitivity analyses (*τ* = 5, *τ* = 10).
- Note that the model assumes two components of infection: contact-based and contact-independent infection. Contact-based infection is proportional to social mixing patterns for France recorded in the COMES-F survey (table S3), and is scaled by the prevalence of severe and mild infection in the age groups that have contact with the focal age group. Contact-independent infection is proportional to the absolute number of severe and mild cases in each age group. The contact-independent term reflects the shedding of virus particles into the environment which may then be acquired as aerosolised particles or through contact with infected surfaces (and for this reason is density rather than frequency-dependent).
- The model is fit to data for consecutive epidemic days *t* = 1.. *T* and for age groups *i* = 1. .8 where the age range for each group is (“0-19”,”20-29”,”30-39”,”40-49”,”50-59”,”60-69”,”70-79”,”>=80”) and where the mean age for each age group is *A*_*i*_ = (9.75, 24.50, 34.58, 44.63, 54.47, 64.42, 73.79, 87.23). The French age distribution that was used was taken from the socialmixr dataset in R ^49^. Day 1, the first day of the dataset, was 01/03/2020. By default *T* = 24 corresponding to the day that we assumed lockdown effects (which commenced on day 17/03/2020 in France) percolated through to new hospital admissions (i.e., 17 + ∼7=∼24 with an assumption of a lower bound of 7 days for time from exposure to hospital admission). Note that these factors are varied in the sensitivity analysis (Fig. S6).
- The model calculates the number of mild cases by dividing the number of severe cases (i.e., numbers in hospital) by the probability of severe disease for the respective age groups, i.e., 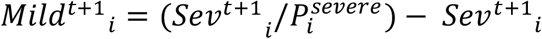. Note that, as we have confirmed using independent simulation, this is a good assumption as long as the epidemic is growing. However, simulations suggest that contrasting removal rates for different age groups and different infection types (i.e., mild vs severe) can lead to a lack of robustness in this assumption beyond the epidemic peak. Note that since we fit up to lockdown the assumption is valid for our purposes.

Note that the model implementation was performed in RStan ^50^ using R. The RStan implementations involved 4 chains, 10,000 iterations (of which 1,000 were warm-up), tree depth=15 and adapt_delta=1-(10^−8^). We obtained the dataset for cumulative age-distributed numbers hospitalised from ‘dailyHospCounts_allReg.csv’ from ^20^. We calculated a dataset of removed (recovered/dead) from the files ‘SIVIC_daily_numbers_region_corrected_histo_20200508.csv’ and ‘SIVIC_total_numbers_region_corrected_histo_20200508.csv’ from ^20^.

##### Model with deviations

The baseline model indicated that an exponential relationship between probability of severe COVID-19 given infection and age is justified. We therefore extended the baseline model to include an assumption of an exponential form and a single key deviation. This model is the basis of Fig. 2 and Table 2 main text (and is reproduced for comparison in Fig. S5A), i.e., we allowed each age-cohort to deviate, denoted *D*_i_, in the probability of severe disease given infection, from the exponential relationship, i.e.

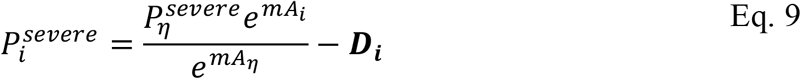

where *A*_*i*_ is the mean age of age group *i* and *A*_η_ is the mean age of the oldest age cohort.

##### Model with further deviations (sensitivity analysis)

In addition we allowed for a further two deviations. These models are the basis of Fig. S5. In the first of the additional model deviations (Fig. S5B), we allowed each age-cohort to deviate in the probability of infection given exposure, denoted *E*_*i*_, from a uniform relationship, i.e.

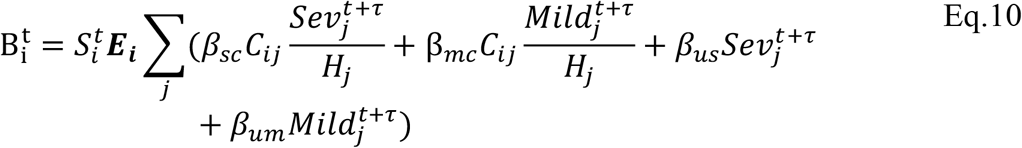

In the second of the additional model deviations (Fig. S5C), we allowed each age-cohort to deviate in the infectiousness of mildly infected individuals, denoted 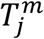 for mildly infected cases, and denoted 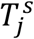 for severely infected cases, from a uniform relationship, i.e.

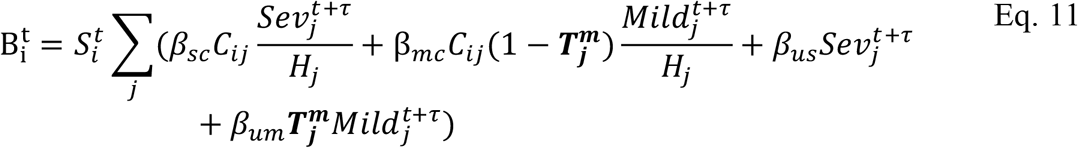

For comprehensiveness, we also considered a variant on the third deviation (Fig. S5D), in which we allowed each age-cohort to deviate in the infectiousness of both mildly and severely infected individuals (denoted 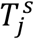 for severely infected cases), i.e.,

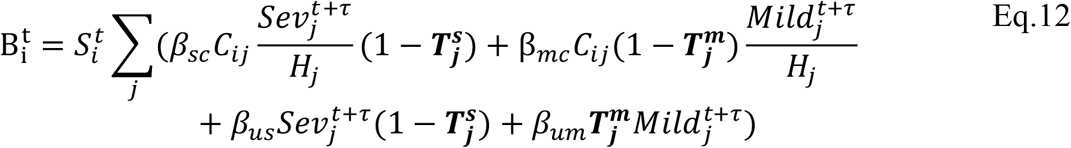

### Data sources

A full list of data sources can be found in Table S3.

Out of the countries we have gathered age-stratified data for, some have reported hospitalisation rates, some all confirmed cases and some both. We have included all hospitalisation data we have collected and only excluded data on all confirmed cases for Italy, since that report came from early in the outbreak, when most, but not all, cases were hospitalisations, therefore giving an exponential relationship with a shallow slope.

For Fig. 1C we used hospitalisation rates and mortality for each age group with non-zero entries for both males and females. We used data from the following countries: France (hospitalisations), England and Wales (all confirmed cases and mortality), England (CHESS program, hospitalisations) and Spain (all confirmed cases, hospitalisations and mortality).

For Fig. S3 we used pre and post lockdown contact matrices from ^20^ which are filtered versions of the contact matrices from socialmixr ^49^.

For the rate of thymus decline, we used the estimate from^5^, which quotes a half-life of 15.7 years, corresponding to an exponential with rate 0.044 years^−1^ or equivalently a rate of 4.5% per year. We used these same numbers when describing the increase in disease risk with age.

We calculated an R^2^ value from the hospitalisation data by focusing on the datasets with the most detailed age-stratification (France, Spain and Denmark) and the age groups with most cases (ages 50-90). Fitting a linear model to the log of the hospitalisation rates gave an exponential rate of 0.046 years^−1^, or equivalently a rate of 4.7% per year with a 95% CI of 4.2-5.2% per year, leading to a 95% CI for the doubling time of 14-17 years. We then fit a linear model with a fixed slope of 0.044 years^−1^, to match the thymic involution timescale, and calculated an R^2^ value of 0.98.

**Extended Data Fig. S1.**
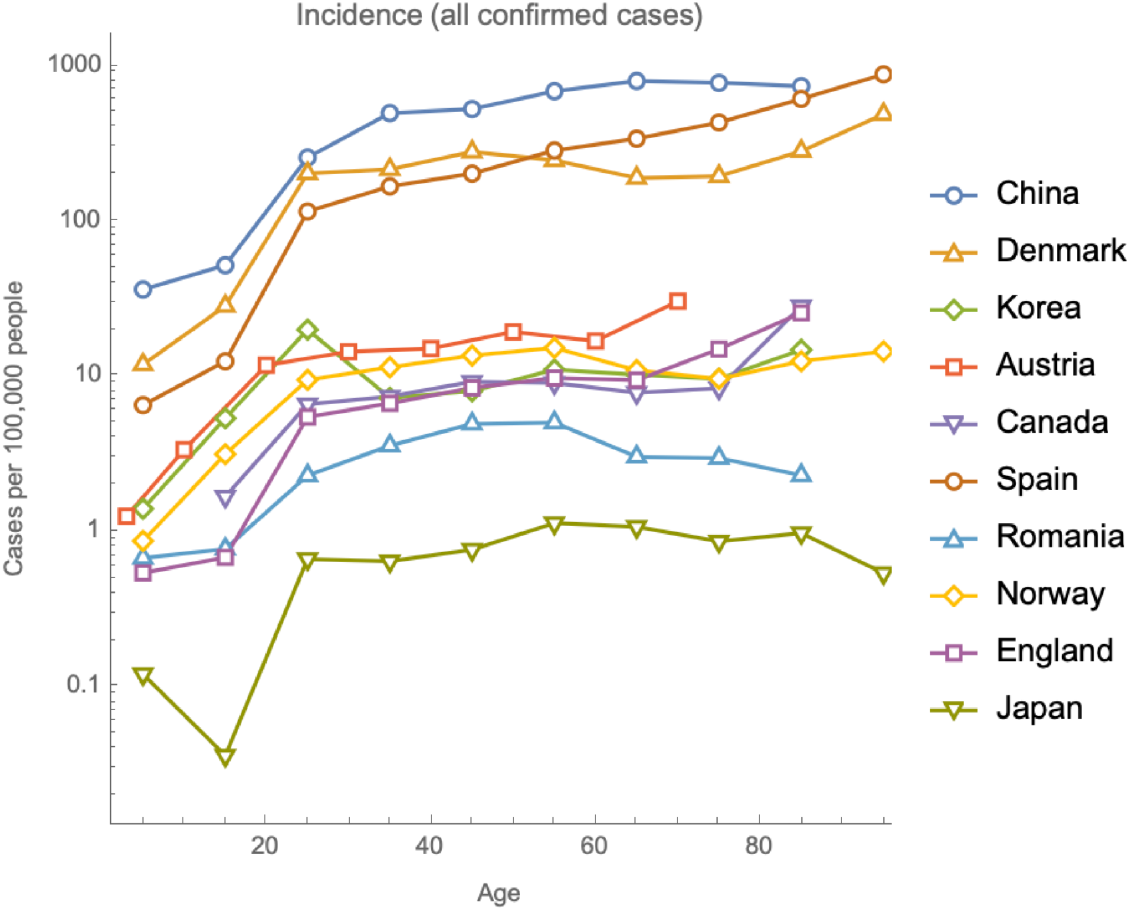
Incidence of all confirmed cases is approximately constant with age for adults and lower in non-adults.

**Extended Data Fig. S2.**
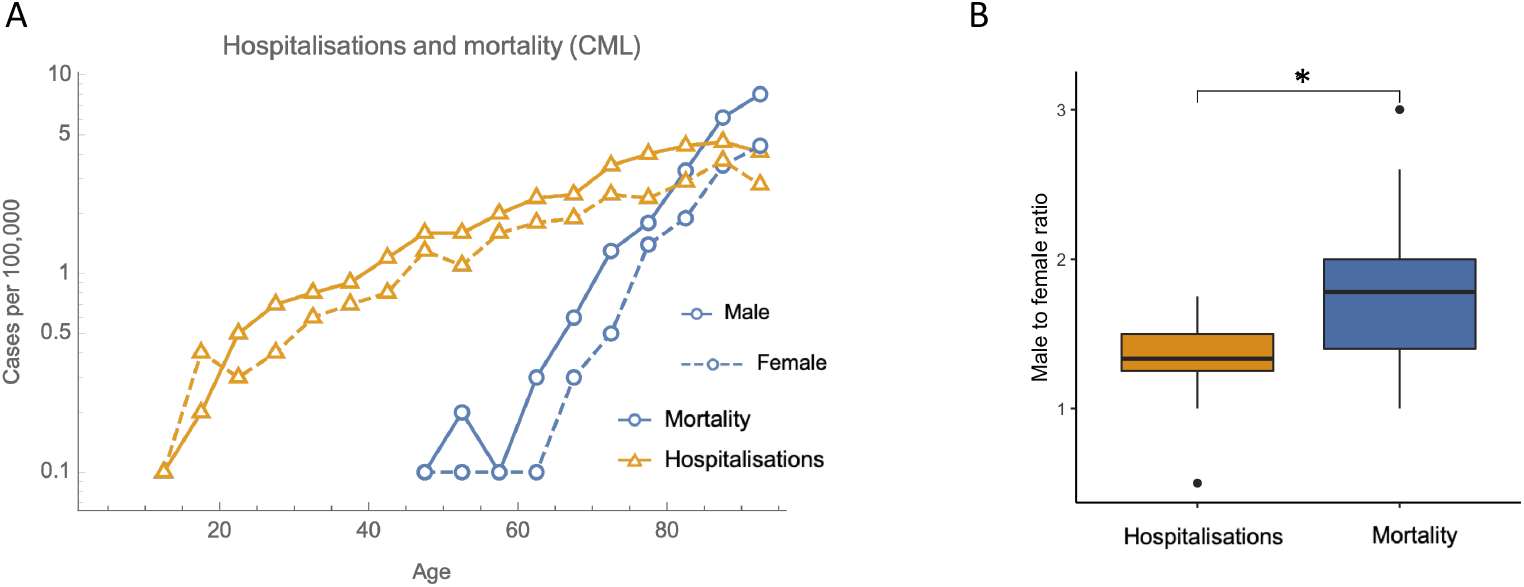
Hospitalisation and mortality rates for CML follow similar trends to COVID-19.

**Extended Data Fig. S3.**
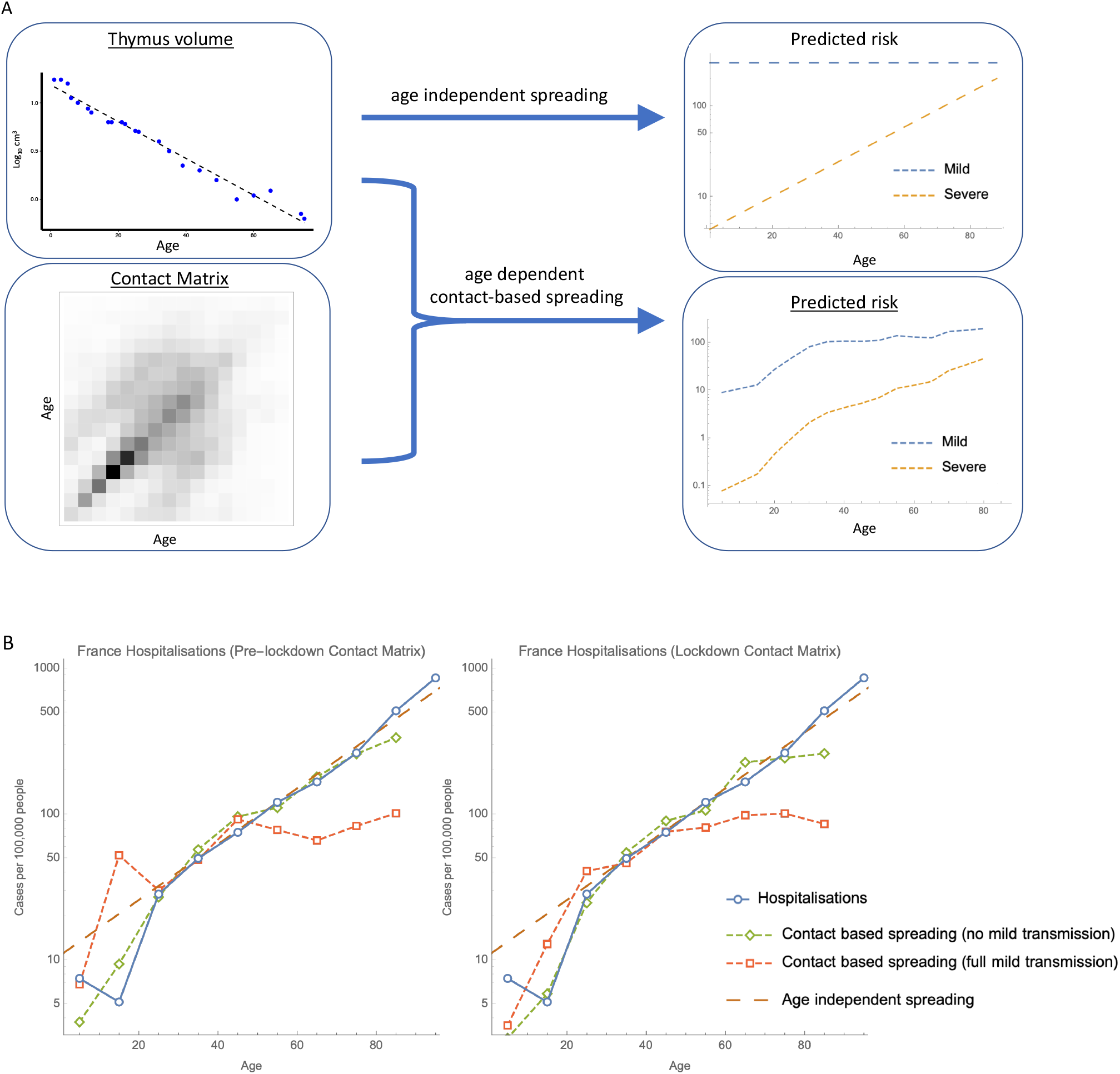
(**A)** Graphical abstract showing the rational for the analytical Markov-chain model (thymus volume data from ref.^5^). If exposure to COVID-19 is age-independent, then a simple model based on immune system declining accurately predicts risk profiles for adults. In this model, the probability of becoming infected is uniform with age and the probability of developing severe symptoms increases exponentially, doubling every 16 years. Alternatively, if the spread of COVID-19 is proportional to contact, as estimated by a contact matrix, then an analytical model can predict similar behaviour for adults and lower risk for non-adults, but only if there is no transmission from mild cases (**B**).

**Extended Data Fig. S4.**
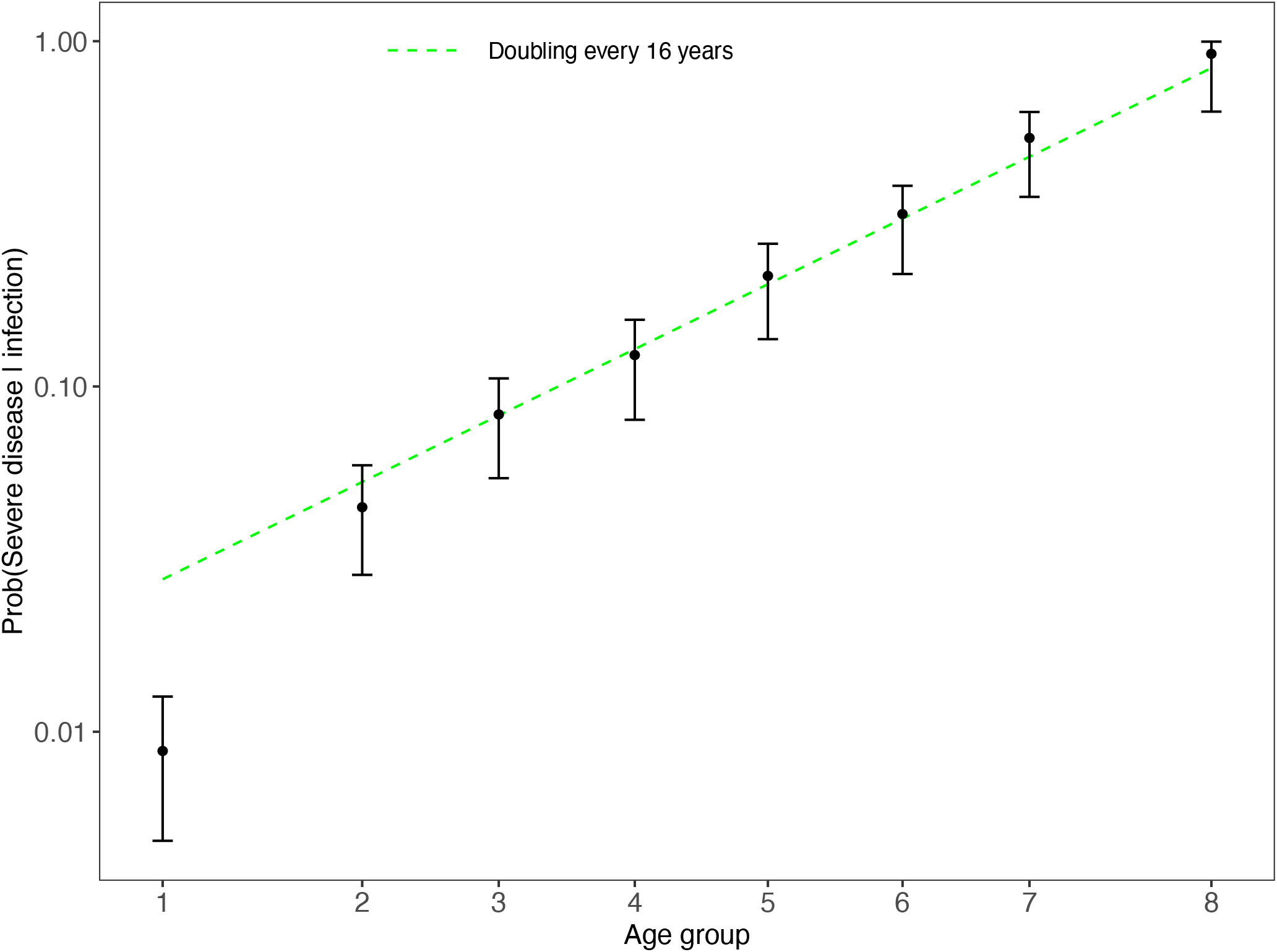
Bayesian analysis of French COVID19 hospitalisation data: the probability that individuals of a given age cohort are hospitalised, given infection, increases exponentially with age. When the probability of hospitalisation given infection was allowed to vary independently for each age cohort, the posterior probabilities of hospitalisation (95% credible intervals in black, with filled circles for mean posterior values) were found to increase at a rate corresponding to thymus degradation for cohorts over the age of 20 (green dashed line) (black dots).

**Extended Data Fig. S5.**
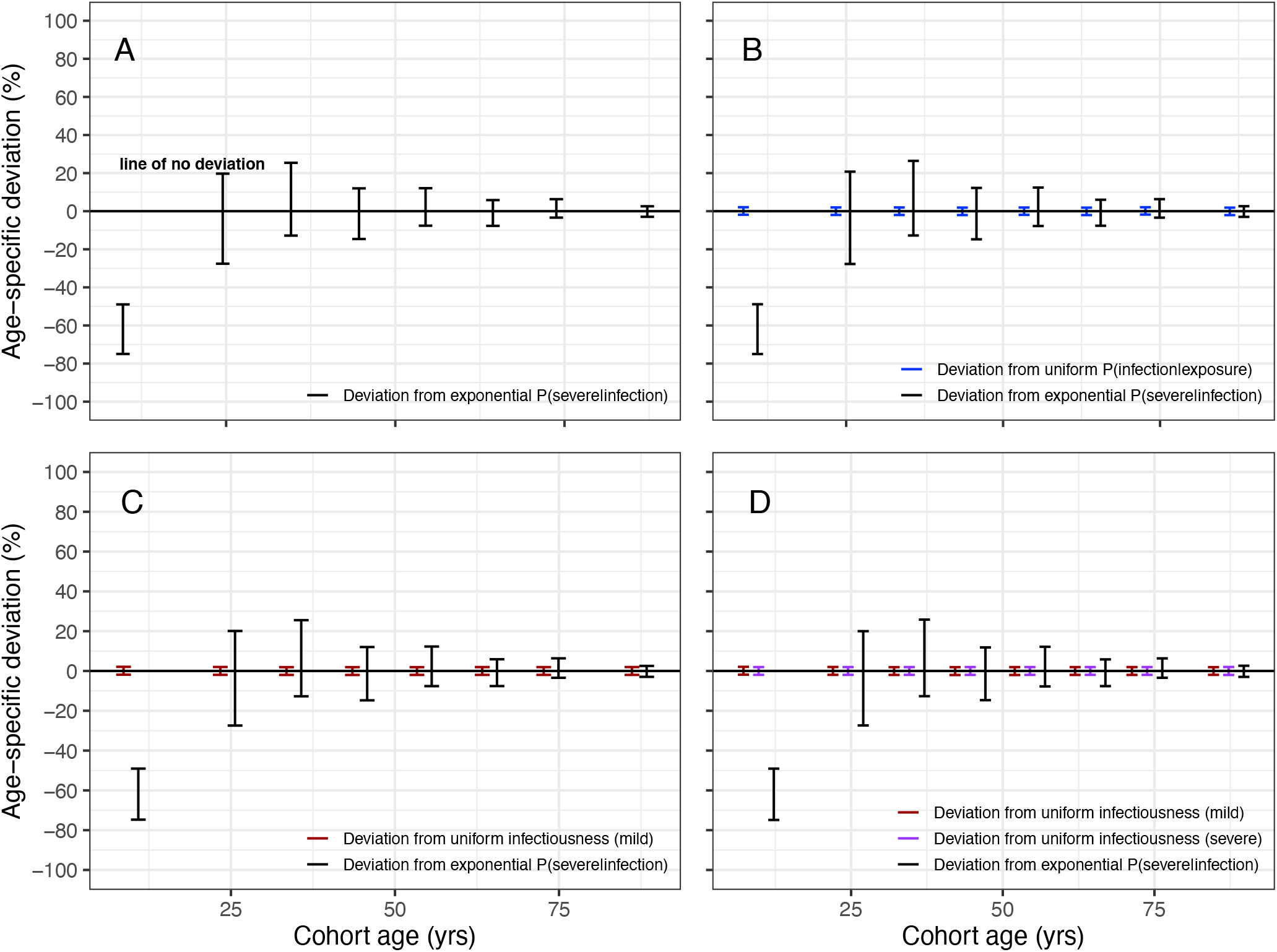
Low COVID-19 hospitalisation among juveniles is associated with additional intrinsic defence beyond strong thymus function instead of age-specific differences in several alternative processes. The likelihood of severe disease increases exponentially with age at a rate matched by thymus degradation, but juveniles deviated significantly from this pattern (A). None of the deviations for the following processes (which may also be explanations for low juvenile severe disease) were significant: age-specific deviations in the probability of infection given exposure (B, blue intervals), or in the infectiousness of mildly infected cases (C, red intervals), or in the infectiousness of mildly and severely infected cases (D, red and purple intervals respectively). The line depicting zero deviation (black, bold, horizontal) passes through each of the credible intervals with the sole exception of the deviation from exponential probability of severe disease for the juvenile age group.

**Extended Data Fig. S6.**
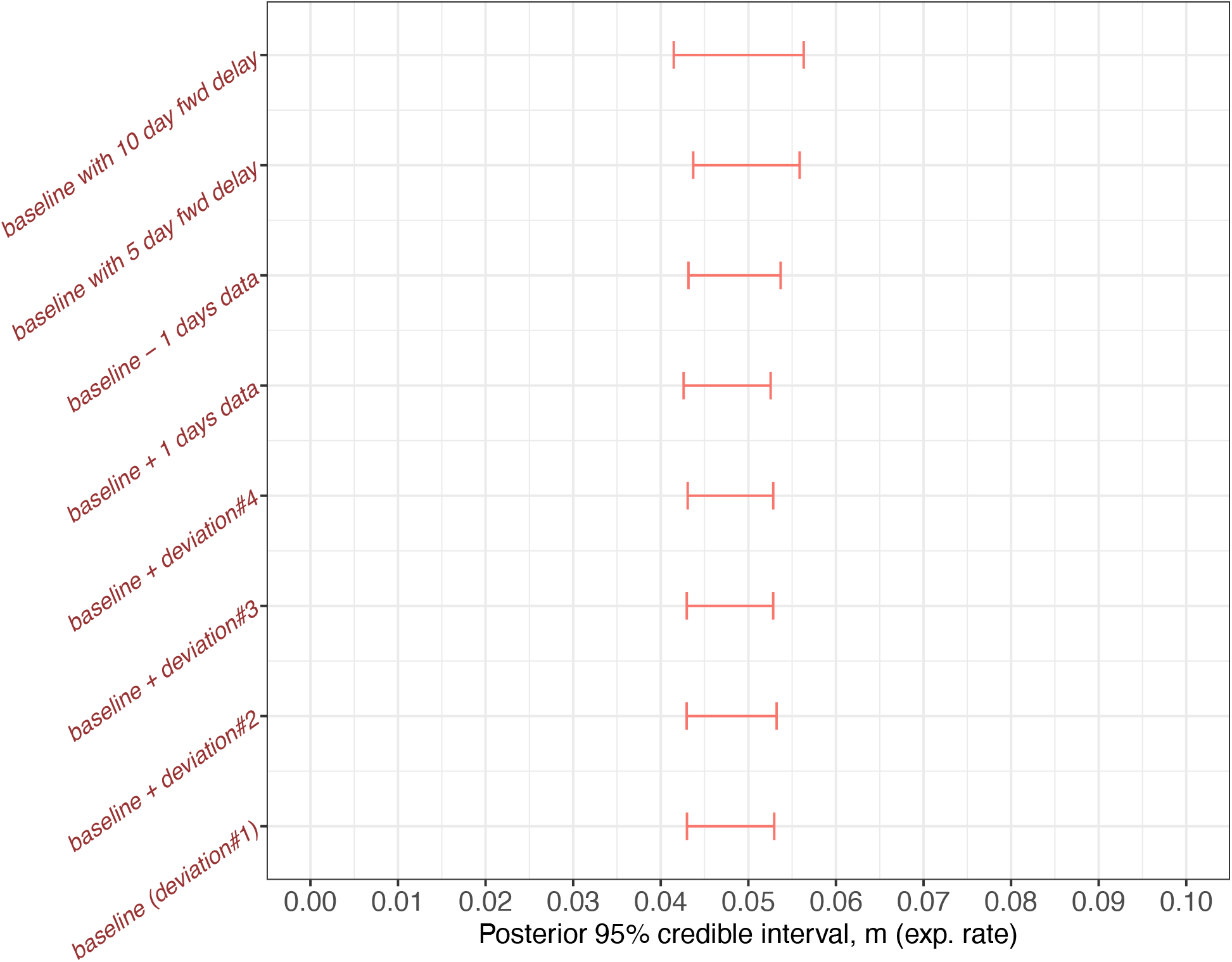
Sensitivity analysis to test the assumptions of the Bayesian analysis of French COVID19 hospitalisation data. Bayesian 95% credible intervals for the rate parameter in the exponential dependence of probability of severe disease. Baseline represents the model in Fig. 2. Baseline+deviation#2 represents the additional inclusion of age-specific deviations from uniform probability of infection given exposure. Baseline+deviation#3 represents the additional inclusion of age-specific deviations from uniform infectiousness of mild cases. Baseline+deviation#4 represents the additional inclusion of age-specific deviations from uniform infectiousness of mild cases and severe cases. Baseline+1 days data represents the extension of the timeseries fit to the model by 1 further day (i.e. baseline model run with 25 days data). Baseline-2 days data represents the truncation of the timeseries fit to the model by 2 days (i.e. baseline model run with 22 days data). Baseline with 5 day fwd delay (and similarly with 10 day fwd delay) represents the baseline model with the rate of occurrence of hospital admissions at time *t* depending on the numbers hospitalised at time *t* + 5 (and similarly depending on the numbers hospitalised at time *t* + 10).

**Extended Data Table S1.** Bayesian 95% credible intervals for epidemiological parameters from French COVID19 hospitalisation data.

| *Epidemiological parameter* | *Mean* | *2.5% (lr 95% threshold)* | *97.5% (upr 95% threshold)* |
| --- | --- | --- | --- |
| $\beta_{sc}/\beta_{mc}$ | 14.89 | 0.04 | 45.11 |
| $\beta_{su}/\beta_{mu}$ | 0.38 | 0.01 | 1.60 |
| $\beta_{sc}/\beta_{su}$ | 9.28 | 0.02 | 38.84 |
| $\beta_{mc}/\beta_{mu}$ | 0.256 | 0.007 | 0.988 |
| $P_{>80y}^{severe}$ | 0.781 | 0.391 | 0.992 |
| *Additional juvenile protection* | 63.4% | 48.9% | 75% |

**Extended Data Table S2.** Parameter definitions, and prior distributions, for the Bayesian analysis. All prior distributions were chosen to be non-informative. Posterior parameter distributions were obtained using Hamiltonian Monte Carlo, RStan version 2.19.3 ^*50*^, R version 3.6.3.

|  | ***Parameters to be fitted*** |  |
| --- | --- | --- |
| $m$ | Probability of severe disease given infection, age coefficient | $\sim \text{gamma}(1,20)$ |
| $\beta_{sc}$ | Contact-based transmission rate from severely infecteds | $\sim \text{gamma}(1,20)$ |
| $\beta_{mc}$ | Contact-based transmission rate from mildly infecteds | $\sim \text{gamma}(1,20)$ |
| $\beta_{su}$ | Contact independent transmission rate from severely infecteds | $\sim \text{gamma}(1,20)$ |
| $\beta_{mu}$ | Contact independent transmission rate from mildly infecteds | $\sim \text{gamma}(1,20)$ |
| $(P_i^{\text{severe}})$ | Inverse of probability of severe disease given infection, age group $i$ | $\sim \text{cauchy}(0, 1)$ |
| $\sigma^{-1}$ | Inverse of negative binomial dispersion exponent | $\sim \text{normal}(2,0.5)$ |
|  | ***Parameters to be fitted, model with deviations*** |  |
| $P_8^{\text{severe}}$ | Prob. of severe disease given infection, for oldest age group $i = 8$ | $\sim \text{cauchy}(0, 1)$ |
| $D_i$ | Deviation from exponential prob. of severe disease given infection, age group $i$ | $\sim \text{normal}(0,0.01)$ |
| $E_i$ | Deviation from uniform prob. of infection given exposure, age group $i$ | $\sim \text{normal}(0,0.01)$ |
| $T_j$ | Deviation from uniform infectiousness of infecteds, age group $i$ | $\sim \text{normal}(0,0.01)$ |
|  | ***Data variables*** |  |
| $H_i$ | Population size, age group $i$, France | *pre-existing data* |
| $Sev_i^t$ | #Hospitalised COVID-19 patients, age group $i$, France | *pre-existing data* |
| $Mild_i^t$ | #Infected with COVID-19 and not hospitalised, age group $i$, France | *Calculated from parameter fits* |
| $R_i^t$ | #Removed, i.e. dead or released, age group $i$, France | *pre-existing data* |
| $C_{ij}$ | #Contacts with age group $j$, age group $i$, France | *pre-existing data* |

**Extended Data Table S3.**
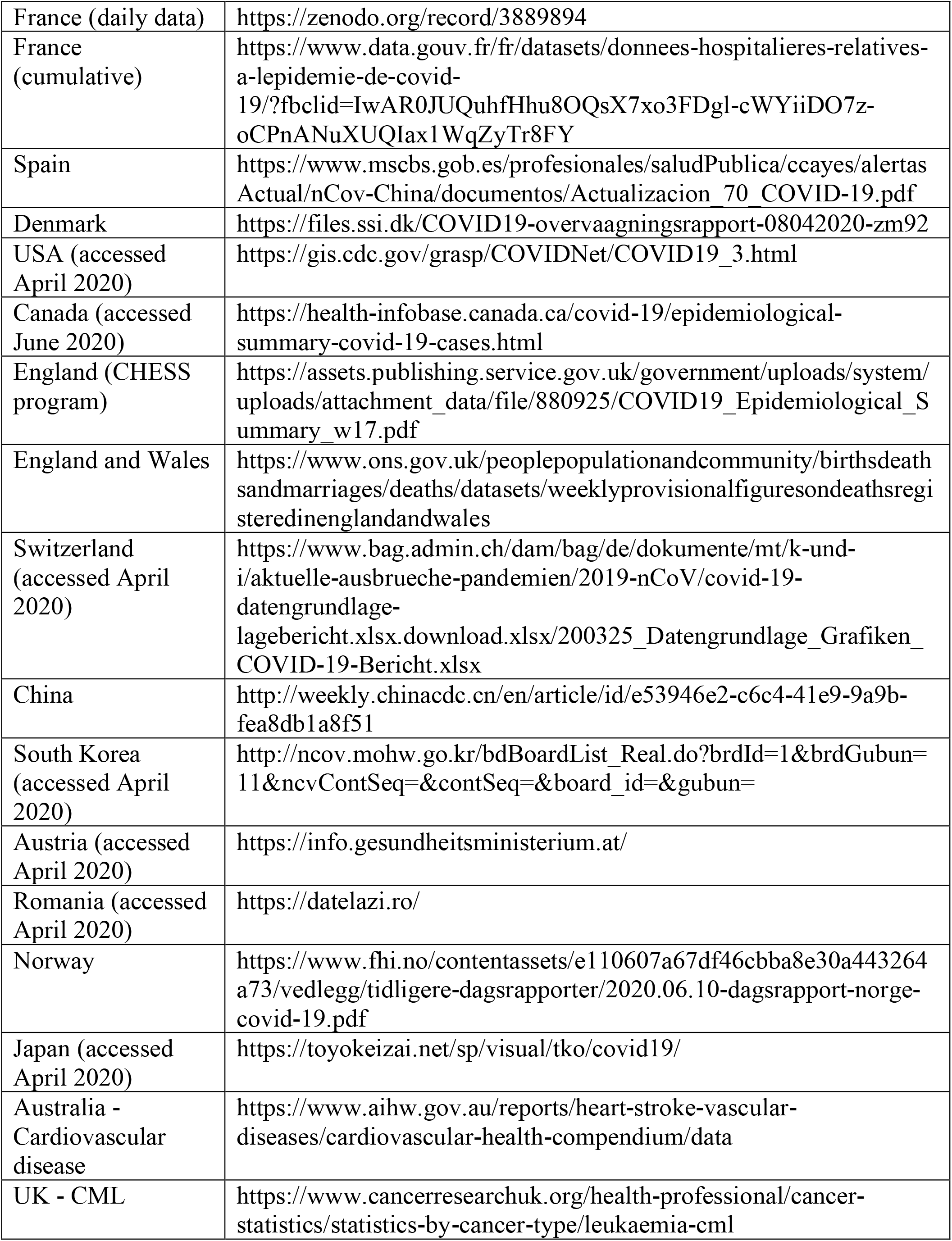
List of data sources for COVID-19 hospitalisations, cardiovascular disease and CML.

